# Underdispersion in the reported Covid-19 case and death numbers may suggest data manipulations

**DOI:** 10.1101/2022.02.11.22270841

**Authors:** Dmitry Kobak

## Abstract

We suggest a statistical test for underdispersion in the reported Covid-19 case and death numbers, compared to the variance expected under the Poisson distribution. Screening all countries in the World Health Organization (WHO) dataset for evidence of underdispersion yields 21 country with statistically significant underdispersion. Most of the countries in this list are known, based on the excess mortality data, to strongly undercount Covid deaths. We argue that Poisson underdispersion provides a simple and useful test to detect reporting anomalies and highlight unreliable data.

## 1 Introduction

Since the beginning of the Covid-19 pandemic, each country in the world has been daily reporting its key Covid statistics, including the number of new Covid cases and the number of new Covid deaths. These numbers are collected into international dashboards, such as the one maintained by the World Health Organization (WHO; https://covid19.who.int), and appear in countless media outlets.

It is, however, well-known that for many countries the reported numbers of cases and deaths can be gross underestimations, due to insufficient testing capacity or in some cases perhaps even purposeful misdiagnosing or misreporting of Covid infections (Kobak, 2021). In some countries the excess mortality, i.e. the increase in the number of all-cause deaths, is much larger than the number of reported Covid deaths, sometimes by over an order of magnitude (Karlinsky and Kobak, 2021), suggesting that the daily reported numbers are unreliable and incomplete.

Here we argue that in some cases the reported numbers exhibit a statistical anomaly, called *underdispersion*, strongly indicative of data tampering and suggesting deliberate obfuscation.

## 2 Results

### 2.1 Underdispersion

Consider the number of Covid-19 deaths reported by the USA during August and September 2021 (Figure 1; here and below we use data distributed by the WHO). This time series shows several prominent features. First, the number of deaths was growing, corresponding to the rising wave of infections. Second, the reported number of deaths had weekly periodicity, with smaller numbers collected during the weekends. Third, on top of the slow monotonic growth and periodic weekly cycle there were additional random fluctuations. Similar patterns are observed in many other countries, which is why these data are typically presented in a smoothed form such as weekly averages.

**Figure 1:**
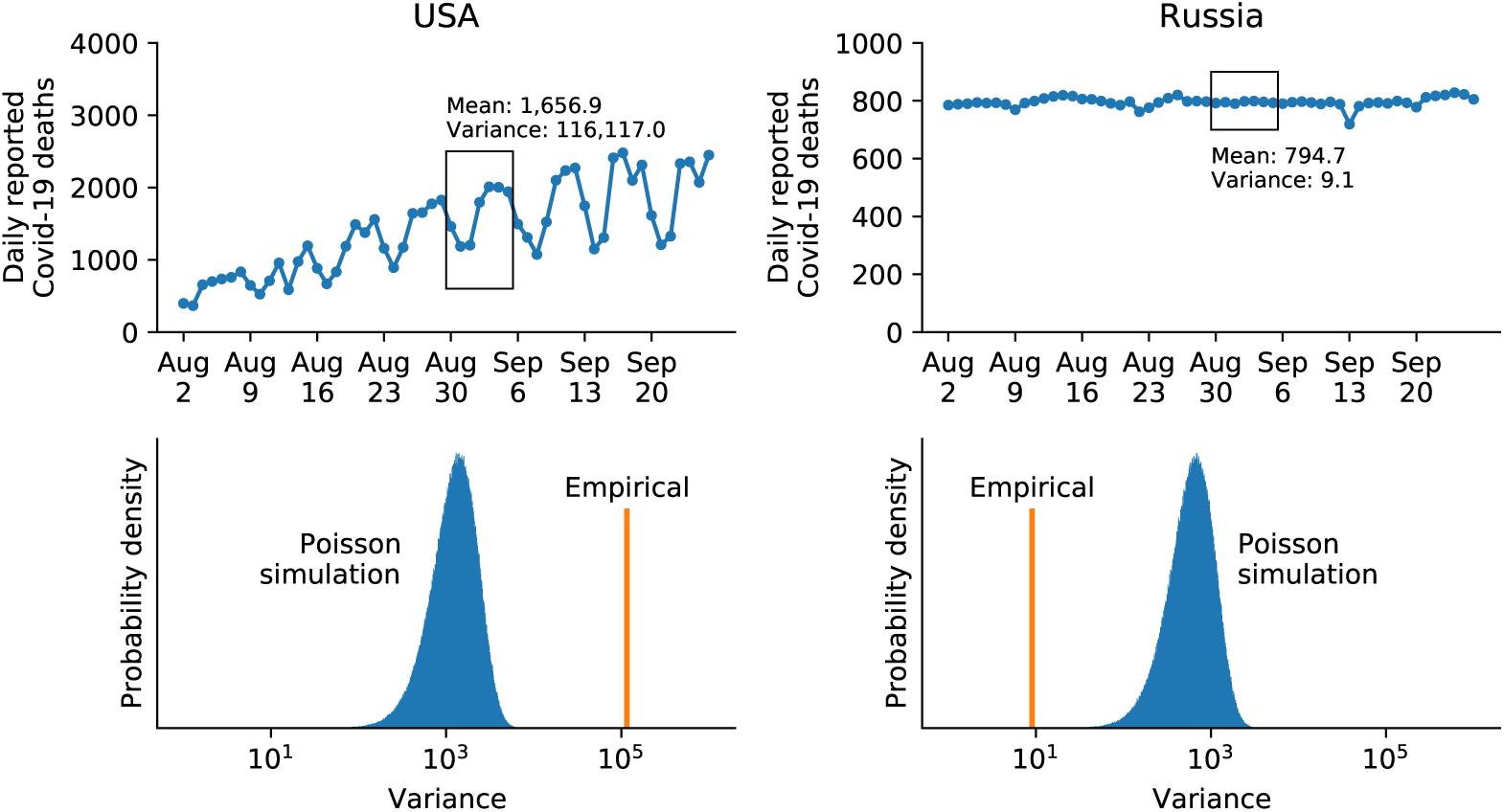
Top left: reported number of daily Covid-19 deaths in the USA over eight weeks in August–September 2021. Ticks on the horizontal axis denote Mondays. The first week of September is highlighted. Top right: the same for Russia. Bottom left: the observed variance of the reported Covid-19 deaths in the first week of September in the USA (orange), and the distribution of variances expected if deaths followed Poisson distribution with the observed mean (blue). Bottom right: the same for Russia.

Some countries, however, did not show any such patterns. For example, the number of reported Covid deaths in Russia over the same time period stayed almost constant at just below 800 deaths per day (Figure 1). Such lack of variation seems inconsistent with the stochastic nature of the data generation process, as people get infected randomly, and the disease progresses independently in each patient. We can get a lower bound on the expected variation using the Poisson distribution. If every citizen in a large country independently had the same small probability of dying from Covid-19 on any given day, daily deaths would follow the Poisson distribution. This remains true if probabilities differ between subpopulations (e.g. depending on geographical region, or person’s sex, or age, etc.), as long as each death is statistically independent. The Poisson distribution has the property that its mean equals its variance, meaning that if 800 people die per day on average, then the Poisson variance would also be 800. In reality the independence assumption does not hold: for example, one superspreading event can lead to an increase in infections or deaths on a particular day, and the observed day-to-day variation will become larger than Poisson. But if the day-to-day variation is smaller than predicted by the Poisson law, it strongly suggests that the time series was not truly random.

We can turn this argument into a formal statistical test. Taking the Russian data from the first week of September (daily deaths: 792, 795, 790, 798, 799, 796, 793; Figure 1, black box), we can compute the average number of daily deaths (794.7) and the observed variance (9.1). The observed variance is clearly much lower than the expected Poisson variance, but it is always possible — if unlikely — that the variance of Poisson random numbers could be as low as this. To find out just how unlikely, we can run a simulation. Simulating 1,000,000 random numbers from the Poisson distribution with the same mean (794.7), we count the number of times the variance turns out smaller than the observed variance (9.1). This happened only 7 times (Figure 1), corresponding to the one-sided *p*-value of 0.000007. The null hypothesis here is that the data are Poisson (or overdispersed), and the alternative hypothesis is that they are underdispersed. The test provides strong evidence that the Russian data were indeed underdispersed. While this conclusion may seem obvious in this particular case, having a formal statistical test will allow us to screen all countries for evidence of underdispersion.

For comparison, we applied the same procedure to the USA data from the same week (daily deaths: 1461, 1185, 1202, 1795, 2010, 2003, 1942; Figure 1, black box). Here the average is 1656.9 and the variance exceeds 115,000 (remember that variance is average squared deviation from the average). Generating 1,000,000 random numbers from the Poisson distribution with the same mean, we never obtained as large a variance (Figure 1), corresponding to the one-sided *p*-value equal to 1. This is not surprising, because the real data are clearly overdispersed compared to Poisson, due to weekly modulation, epidemic growth, and other possible sources of variability. Note that the USA reports Covid-19 deaths by date of registration, but we observed Poisson overdispersion in many other countries that report Covid-19 deaths by date of death (and thus do not show weekly modulation), such as Belgium, Spain, and Sweden.

### 2.2 Screening for underdispersion

With this statistical tool at hand, we tested for underdispersion all officially reported Covid-19 cases and deaths time series in the WHO dataset. This dataset contains data on 237 countries and territories, and we analyzed each week (from Monday to Sunday) between 3 March, 2020, and 30 January, 2022 (100 weeks in total). In each week, we took underdispersion as statistically significant, whenever we obtained *p ≤* 0.05 (using 1000 Poisson simulations). We have thus conducted 237 100 2 separate statistical tests. To mitigate the multiple testing problem, we considered a country showing statistically significant underdispersion if it either had at least 15 weeks with statistically significant underdispersion, or at least 4 weeks in a row. The probability of that happening under the null hypothesis is 0.0007, so even when testing 237 countries, we should expect to see almost no false positives.

Seventeen countries came out with statistically significant underdispersion in the Covid-19 deaths, and 3 more countries in Covid-19 cases (Figure 2). As we discuss below, for many of these countries, there is strong independent evidence that Covid-19 deaths have been undercounted, suggesting that the underdispersion test yielded a sensible list of countries with suspicious Covid reporting.

**Figure 2:**
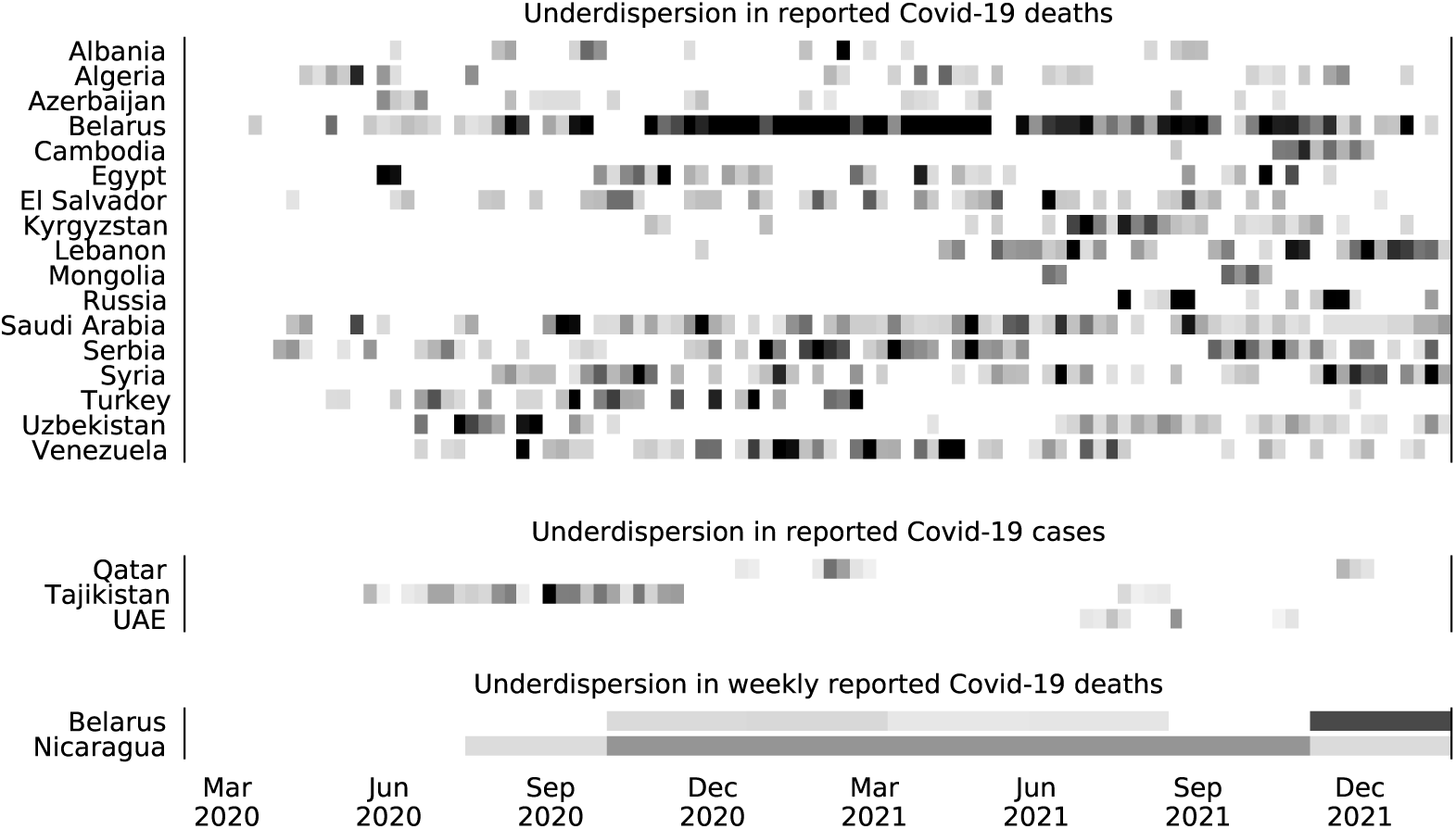
Screening all 237 countries and territories in the WHO dataset for underdispersion in daily reported Covid-19 deaths and cases numbers between 3 March, 2020, and 30 January, 2022 (100 weeks). Gray rectangles denote weeks (from Monday to Sunday) showing statistically significant (*p ≤* 0.05) underdispersion compared to the Poisson distribution with the same mean. Only countries with at least 15 significant weeks in total or at least 4 significant weeks in a row are shown. Shades of gray denote the mean divided by the variance in each week (black: 20 or higher). Bottom: screening for underdispersion in weekly reported Covid-19 deaths in blocks of 10 weeks. Only countries with at least 5 significant blocks are shown.

We additionally tested all 85 Russian federal regions as well as all 60 public health jurisdictions in the USA. In Russia, 82 regions out of 85 were flagged for underdispersion, with many regions showing statistically significant underdispersion during long periods of time, much longer than twelve weeks that we obtained for Russia as a whole (Figure 2). This suggests that data tampering occuring at regional level may become invisible at the country level, and also that a separate mechanism may have been implemented in Russia in August-September 2021 to maintain the number of reported deaths just below 800 on the federal level. In contrast, for USA jurisdictions, not a single one showed underdispersion.

Furthermore, we tested reported Covid-19 numbers for underdispersion across weeks. We summed reported Covid-19 cases and deaths within each week, and applied the same procedure as above to test for underdispersion of weekly counts over windows of ten consecutive weeks (100/10 = 10 windows in total). Only two countries showed statistically significant underdispersion in at least five windows (probability of this happening under the null hypothesis was below 0.0001): Belarus and Nicaragua (Figure 2). Nicaragua is a particularly telling case as it has been reporting exactly 1 death per week since the beginning of 2021. This clearly implausible lack of variation is correctly picked up by our test.

### 2.3 Relationship to excess mortality

We have previously defined the undercount ratio as the ratio of the excess deaths during the pandemic to the officially reported Covid deaths (Karlinsky and Kobak, 2021). For countries with reliable reporting, this ratio is around 1.0 or often even below 1.0. But for many countries listed in Figure 2 the undercount ratio was much larger than that: at the time of writing (January 31, 2022), it is estimated to be 4.1 in Albania, 18.3 in Algeria, 4.3 in Azerbaijan, 14.4 in Belarus, 12.9 in Egypt, 6.6 in El Salvador, 4.9 in Kyrgyzstan, 50.1 in Nicaragua, 3.5 in Russia, 4.0 in Serbia, 23.4 in Uzbekistan, and 104.7 in Tajikistan (https://github.com/dkobak/excess-mortality). For Turkey, the undercount ratio has been estimated at *∼*3, based on incomplete excess mortality data (https://gucluyaman.com/excess-mortality-in-turkey). For Syria, the undercount ratio in its capital Damascus has been estimated at *∼*17, based on obituary notifications (Watson et al., 2021). Large undercount ratios suggest purposeful misreporting or misdiagnosing of Covid deaths (Karlinsky and Kobak, 2021), in good agreement with the idea that underdispersion may also be indicative of data manipulations.

Only three countries flagged for underdispersion in Figure 2 have moderate undercount ratios, namely Lebanon (1.5), Mongolia (<1), and Qatar (1.5). For Cambodia, Saudi Arabia, United Arab Emirates (UAE), and Venezuela, no data on excess deaths are available so far. Our results suggest that the undercount there may be large.

Overall, 8 out of 10 countries with the highest undercount ratios in the World Mortality Dataset (Karlinsky and Kobak, 2021) at the moment of writing demonstrated statistically significant underdispersion in our analysis. To quantify the relationship between undercount and underdispersion, we defined the underdispersion index as the ratio of the observed mean to the observed variance (averaged over 100 weeks or 10 ten-week windows; for each country we took the maximum over four such average values, obtained for daily and weekly numbers of deaths and cases). The correlation of underdispersion index to undercount ratio was 0.40 (Figure 3). Most importantly, large values of the underdispersion index were always associated with high undercount. This suggests a large undercount in several countries with large values of the underdispersion index but no currently available estimates of excess mortality (e.g. Saudi Arabia, Syria, Turkey, Venezuela).

**Figure 3:**
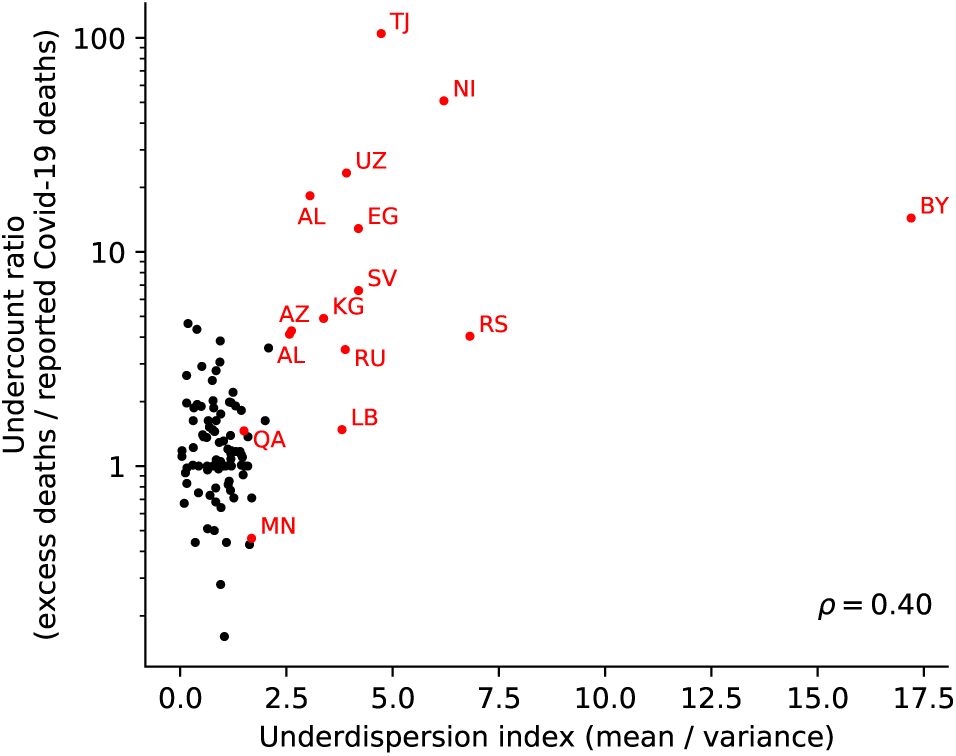
Relationship between the underdispersion index (ratio of observed mean to observed variance; averaged over 100 weeks or 10 ten-week windows; maximum over four such average values, obtained for daily and weekly numbers of deaths and cases; we added 0.1 to the observed variances to avoid division by zero) and the undercount ratio (ratio of excess deaths to the reported deaths; for countries with negative or statistically not significant excess mortality we here set the undercount ratio to 1) (Karlinsky and Kobak, 2021). Pearson correlation 0.40 (*n* = 113; all countries present in both, the World Mortality Dataset and the WHO dataset). Red dots denote countries listed in Figure 2, labeled with their ISO codes. Note that many countries do not have any estimate of the undercount ratio due to lack of excess mortality data; those are not shown.

## 3 Discussion

Underdispersion in the reported Covid-19 cases or deaths numbers cannot serve as a definite proof of data tampering. Underdispersion can in principle arise through other mechanisms, such as e.g. a country reaching a limit of its testing capacity and so reporting the same, maximally possible, number of new cases every day. We think that this is unlikely to explain the observed patterns: most of the underdispersion was observed in the reported deaths while the number of reported cases was much larger (so Covid tests could not have been the limiting factor). Also, in many cases, underdispersion was observed during weeks with reported numbers smaller than reported by the same country in the past, again suggesting that testing or reporting capacity could not have been reached.

We believe that the most likely explanation for observed underdispersion patterns is deliberate data tampering, when a country or a region did not report the same values as were in fact internally obtained. Such tampering may not necessarily have malicious intent (e.g. if somebody was ‘re-distributing’ the values across days), but, together with the evidence of underreporting coming from excess mortality, is strongly suggestive of it.

Importantly, if a country did not show evidence of underdispersion, it does not mean that Covid reporting there has been accurate. There may exist other numerical anomalies indicative of misreporting, beyond the underdispersion. Furthermore, there is strong evidence that in many developing countries without reliable mortality tracking the Covid undercount may be very high (Whittaker et al., 2021), even though the reporting itself could be honest and without any statistical anomalies. Despite these obvious limitations, here we argued that Poisson underdispersion provides a simple and useful test to detect one kind of reporting anomalies and highlight unreliable data.

## Data Availability

All data produced are available online at https://github.com/dkobak/covid-underdispersion.

## Code availability

Analysis code together with the frozen data is available at https://github.com/dkobak/covid-underdispersion.

## Acknowledgments

The author thanks Ariel Karlinsky for comments and suggestions, and Sergey Shpilkin for scraping and sharing the regional Russian data.

